# Immediate post-operative PDE5i Therapy improves early Erectile Function Outcomes after Robot Assisted Radical Prostatectomy (RARP)

**DOI:** 10.1101/2020.09.25.20200683

**Authors:** A. Nathan, S. Shukla, A. Sinha, S. Sivathasan, A. Rashid, J. Rassam, S. Smart, K. Patel, N. Shah, B.W. Lamb

**Affiliations:** Addenbrooke’s Hospital, Cambridge; University of Cambridge

**Keywords:** Potency, Erectile Dysfunction, Phosphodiesterase Inhibitors (PDE5i), Robot-Assisted Radical Prostatectomy (RARP), Continence

## Abstract

**Objectives:** To assess whether the timing of post-operative Phosphodiesterase Inhibitor (PDE5i) therapy after Robot Assisted Radical Prostatectomy (RARP) is associated with a change in early erectile function outcomes. Additionally, to determine whether there are differences in continence or safety outcomes.

**Subjects/patients and methods:** Data was prospectively collected from a single surgeon in one tertiary centre and retrospectively evaluated. 158 patients were treated with PDE5i therapy post RARP over a two-year period. PDE5i therapy was started: immediately (day 1-2) post-op in 29%, early (day 3-14) post-op in 37% and late (after day 14) post-op in 34%. EPIC-26 Erectile Function (EF) scores were collected pre-op and post-op with a median follow-up time of 43 days.

**Results:** The median age was 64 and the median BMI was 27. 9% of the series had Charlson Co-Morbidities. There were no significant differences in pre-operative characteristics between the therapy groups.

Patients that had bilateral nerve sparing had a mean drop in Erectile Function (EF) score by 5.4 compared to 8.8 in the unilateral group. Additionally, 34.9% of bilateral nerve sparing patients returned to baseline compared to 12.1% of unilateral.

Drop in EF scores and percentage return to baseline for unilateral nerve sparing was respectively 9 and 11.1% of immediate therapy, 7 and 14.8% of early therapy and 9.7 and 9.5% of late therapy (p=0.9 and p=0.6). For bilateral nerve sparing this was respectively 3.5 and 42.9% immediate therapy, 5.5 and 35.5% early therapy and 7.3 and 25% late therapy (p=0.017 and p=0.045).

Pad free and social continence was achieved in 54% and 37% of those receiving immediate therapy, 60% and 33% for early therapy and 26% and 54% for late therapy. There were no differences in compliance, complication or readmission outcomes.

**Conclusion:** In patients with bilateral nerve sparing RARP, immediate post-operative PDE5i therapy may protect EF. Early onset (3-14 days) may also provide a benefit compared to initiating PDE5i therapy later (after 14 days). Expediting therapy for patients undergoing unilateral nerve sparing may also provide a benefit; however, the differences are less pronounced. Immediate or early PDE5i therapy also improved early continence outcomes for patients with bilateral nerve sparing, compared to late therapy. There were no differences in compliance, complication or readmission rates between the groups. Therefore, immediate PDE5i therapy should be considered in patients following nerve sparing RARP in order to maximise functional outcomes, especially in those undergoing bilateral nerve spare.

## Introduction

Prostate Cancer (PCa) is the most commonly diagnosed cancer in men within the UK, accounting for 26% of all oncological diagnoses in 2017 (1). The 2019 National Prostate Cancer Audit (NPCA) reported that 33% of 21,896 men treated for PCa underwent radical prostatectomy (2). Robot Assisted Radical Prostatectomy (RARP) is a minimally invasive procedure which has been rapidly adopted by urological surgeons worldwide (3). Despite the learning curve and costs involved, RARP has shown equal or better cancer and quality outcomes compared to open surgery (4-6).

Despite the benefits of RARP, post-operative Erectile Dysfunction (ED) continues to be an issue for patients. A meta-analysis of potency rates by Ficcara *et al* (7) reported the incidence of ED in men undergoing either unilateral or bilateral nerve sparing to be 68%, 47%, 31% and 37% at 3, 6, 12 and 24 month follow-up intervals. When considering bilateral nerve-sparing only the ED rates were better at 44%, 31%, 26%, and 18% respectively. Return to baseline function takes approximately six months to two years (8). Furthermore, early potency rates post RARP can be as low as 33% during the six-month follow-up period (9). Salvage RARP has shown inferior Erectile Function (EF) outcomes compared to primary RARP with only 5% maintaining their pre-operative EF with two-year follow-up (10).

Crucially, EF has been shown to have significant effects on the psychological wellbeing of patients with studies showing over half of patients (51.6%) can feel angry, bitter or depressed at the loss of potency (11, 12).

As well as being affected by age, BMI, co-morbidities, baseline erectile function and nerve-sparing, EF outcomes are also affected by the use of penile rehabilitation therapy following surgery (13).

The use of phosphodiesterase inhibitors (PDE5i) following RARP, as part of penile rehabilitation, has been shown to preserve EF. ED post RARP can be explained by intra-operative damage to the pelvic splanchnic nerves. Upon sexual stimulation the pelvic splanchnic nerves release Nitric Oxide (NO) from Non-Noradrenergic Non-Cholinergic (NANC) fibres and Acetylcholine (ACh) from parasympathetic fibres leading to an overall increase in cyclic-GMP (cGMP) concentration. Subsequently there is a reduction in calcium ion concentration leading to the relaxation of vascular smooth muscle. Laboratory studies have demonstrated that PDE5i’s block the action of phosphodiesterase which acts to break down cGMP, thereby working to reduce calcium ion concentration and promoting relaxation of vascular smooth muscle (14, 15).

Furthermore, numerous clinical studies have shown that the use of PDE5i’s following RARP improve EF for patients with nerve-sparing surgery. Briganti *et al* followed patients treated by high volume surgeons with bilateral nerve sparing over a period of three-years. They found EF recovery rates to be significantly higher (73%) in the PDE5i therapy group compared to the placebo (37%) group (16). Similarly, Nelson *et al* found the EF recovery rate in the PDE5i group to be 43% compared to the placebo group of 22% at two-years post-op (17). Mulhall *et al* found that three months of on-demand 200mg Avanafil treatment significantly improved EF, with outcomes of 41% in the treatment group and 10.7% in the placebo group (18). Current favourable evidence is better established for bilateral nerve sparing compared to unilateral nerve sparing (19, 20). The limited evidence for PDE5i therapy for patients with no nerve sparing shows no benefit (21).

Despite considerable evidence supporting the efficacy of PDE5i therapy on EF post RARP, there are few guidelines or evidence-based recommendations advising the optimal time to begin PDE5i therapy to begin post RARP. Teloken *et al* questioned 301 doctors from 41 countries and found 95% use PDE5i therapy post RARP however only 6% initiate therapy immediately post-operatively. 54% waited until after the catheter removal around two weeks and the remaining 40% all started therapy at some point in the first four months (22). Moreover, some studies have suggested that the early use of PDE5i may be associated with an increased incidence of adverse events and worsen early continence outcomes (23, 24).

## Aims

We aim to assess if there is an association between the timing of initiation of PDE5i therapy and early EF outcomes. Additionally, we aim to assess if the timing of initiation of PDE5i therapy is associated with adverse events or if it affects continence outcomes.

## Subjects/Patients and Methods

Data was prospectively collected from a single surgeon at one tertiary centre. Data was then retrospectively evaluated. All patients were registered as part of The BAUS national outcomes audit and registered with the institutional audit department (Ref. PRN8750). 187 patients who underwent primary RARP were identified over a two-year period from 2018 to 2020, from these 158 patients were treated with PDE5i therapy. 29 patients did not receive PDE5i therapy due to non-nerve sparing status or lack of patient interest and were excluded.

The decision to attempt nerve sparing surgery was made jointly between the operating surgeon and patient based on a synthesis of patient priorities and disease factors. Specifically, patient priorities included the relative prioritisation of functional recovery and oncological control. Disease factors included the location lesion on the MRI scan, PSA level, as well as the distribution and grade of biopsy cores with PCa. Finally, digital rectal examination under anaesthesia was performed at the start of the case. The definition of nerve sparing included intrafascial and interfascial, as well as high- and low-release of the neurovascular bundle. The execution of nerve sparing reflected the intent as set out above, as well as what was intraoperatively feasible. All patients gave informed consent for surgery and PDE5i therapy. All patients were included in the analysis regardless of baseline EF or nerve-sparing status.

Three cohorts were stratified based on the time PDE5i therapy was initiated post RARP. Tadalafil 5mg once daily was the PDE5i of choice. In 2018, patients were started on late therapy however due to emerging evidence, practice was changed to starting therapy immediately post-op (25-27). Furthermore, due to anecdotal concerns regarding safety, initiation time was delayed to early post-operative therapy (28). Some patients received specific treatment depending on individual risk factors such as nerve-sparing status and risk of haematoma.

EF and post-operative continence were measured using the patient reported outcome measure (PROM) - The Expanded Prostate Cancer Index Composite Short Form (EPIC-26). EPIC −26 is a questionnaire designed to measure Quality of Life issues in patients with PCa. EF was measured using questions 8-11 with a minimum score of 5/24 and a maximum score of 24/24 (29).

Pre-operative metrics such as age, BMI, Charlson Co-Morbidities and intra-operative nerve spare status; as well as post-operative data on continence, complications, readmission and discontinuation were also collected.

Return to Baseline was defined as the post-operative EF score equalling or exceeding the pre-operative EF score (30). Full continence was defined as using zero pads per day, social continence was defined as using one pad per day and incontinence was defined as using two or more pads per day (31, 32).

Data collection, tables and figures were completed using Microsoft Excel 2019. Statistical analysis was done using SPSS 26^th^ Edition, IBM. Chi-Squared tests were used to compare categorical data and independent T-tests and ANOVA were used to compare continuous data.

## Results

### Demographics

For the entire cohort, median age was 64 (IQR: 59-67); median BMI was 27 (25-30); Charlson Co-Morbidities were present in 9%. Forty-six (29%) patients received PDE5i therapy immediately between day 1-2 (median = day 1) post-operatively. 58 (37%) patients received PDE5i therapy early between day 3-14 (median = day 10) post-operatively. 54 (34%) patients received PDE5i therapy late after day 14 (median = day 40). There were no statistically significant differences in age, BMI, Co-Morbidities or pre-operative EF scores between the three groups. 53% of our cohort had bilateral nerve-sparing, 42% had unilateral nerve-sparing and 6% had no nerve-sparing (Table 1). The median follow-up time was 43 days (IQR: 41-47).

**Table. 1.** Pre-operative and intraoperative characteristics of therapy groups by PDE5i therapy onset.

|  | Total | Immediate | Early | Late | P-Value |
| --- | --- | --- | --- | --- | --- |
| **Criteria (Day Post-Op)** |  | 1-2 | 3-14 | >14 |  |
| **N (%)** | 158 (100) | 46 (29) | 58 (37) | 54 (34) |  |
| **Pre-Op Demographics** |  |  |  |  |  |
| **Median Age (IQR) years** | 64 (59-67) | 66 (62-68) | 62 (57-66) | 64 (59-67) | 0.916 |
| **Median BMI (IQR)** | 27 (25-30) | 28 (25-30) | 27 (25-30) | 27 (24-29) | 0.854 |
| **Co-Morbidities N (%)** | 14 (9) | 3 (7) | 4 (7) | 7 (13) | 0.424 |
| **Nerve-Sparing Status** |  |  |  |  |  |
| **Nil N (%)** | 9 (6) | 0 (0) | 0 (0) | 9 (6) | 0.210 |
| **Unilateral N (%)** | 66 (42) | 18 (11) | 27 (17) | 21 (13) |  |
| **Bilateral N (%)** | 83 (53) | 28 (18) | 31 (20) | 24 (15) |  |

N = Numbers, \* = statistically significant at p<0.05

### Erectile Function Outcomes

There was a mean drop in EF score of 9.4 and return to baseline of 22% for no nerve sparing. Unilateral nerve sparing achieved 8.8 and 12% whilst bilateral achieved 5.4 and 35% (p<0.05) (Table 2).

**Table. 2.**
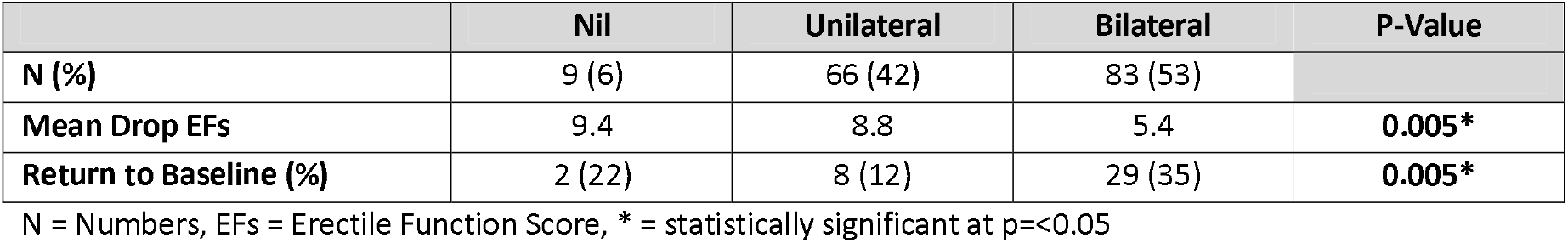
Erectile Function Outcomes between nerve-sparing groups.

The mean pre-operative EF score was 17.4 out of 24, mean drop in EF score was 7 and the return to baseline was 25%. Mean drop in EF and return to baseline was 5.7 and 36% with immediate therapy, 6.6 and 39% with early therapy and 8.6 and 26% with late therapy (p=0.097 and p=0.374) (Table 3).

**Table. 3.** Post-operative outcomes of therapy groups by PDE5i therapy onset.

|  | Total | Immediate | Early | Late | P-Value |
| --- | --- | --- | --- | --- | --- |
| **N (%)** | 158 (100) | 46 (29) | 58 (37) | 54 (34) |  |
| **Erectile Function** |  |  |  |  |  |
| **Mean Pre-Op EFs** | 17.4 | 17.9 | 18.2 | 16.2 | 0.110 |
| **Mean Drop EF** | 7 | 5.7 | 6.6 | 8.6 | 0.097 |
| **Return to Baseline N (%)** | 39 (25) | 14 (36) | 15 (39) | 10 (26) | 0.374 |
| **Continence** |  |  |  |  |  |
| **Full Continence N (%)** | 74 (47) | 25 (54) | 35 (60) | 14 (26) | **0.001\*** |
| **Social Continence N (%)** | 65 (41) | 17 (37) | 19 (33) | 29 (54) | **0.043\*** |
| **Incontinence N (%)** | 19 (12) | 4 (9) | 4 (7) | 11 (20) | 0.065 |
| **Safety** |  |  |  |  |  |
| **Stopped N (%)** | 21 (13) | 6 (13) | 3 (5) | 12 (22) | 0.589 |
| **-Due to Side Effects N (%)** | 11 (7) | 5 (11) | 2 (3) | 4 (7) | 0.934 |
| **-Due to Compliance (%)** | 10 (6) | 1 (2) | 1 (2) | 8 (15) | 0.574 |
| **Complications N (%)** | 8 (5) | 2 (4) | 2 (3) | 4 (7) | 0.948 |
| **Readmissions N (%)** | 3 (2) | 1 (2) | 1 (2) | 1 (2) | 0.865 |

N = Numbers, EFs = Erectile Function Score, \* = statistically significant at p<0.05
Full continence = 0 pads, Social continence = 1 pad / day, Incontinence = 2 or more pads / day

For unilateral nerve spare, mean drop in EF score and return to baseline was 9 and 11% for immediate, 7 and 15% for early and 9.7 and 10% for late. For bilateral nerve spare, mean drop in EF score and return to baseline was 3.5 and 43% for immediate, 5.5 and 36% for early and 7.3 and 25% for late (p<0.05) (Table 4) (Fig 1) (Fig 2).

**Table. 4.** Erectile Function outcomes by nerve spare and PDE5i therapy onset.

|  | Total | Immediate |  |  | Early |  |  | Late |  |  |
| --- | --- | --- | --- | --- | --- | --- | --- | --- | --- | --- |
| Nerve-sparing |  | Nil | Uni | Bi | Nil | Uni | Bi | Nil | Uni | Bi |
| N | 158 | 0 | 18 | 28 | 0 | 27 | 31 | 9 | 21 | 24 |
| Mean Pre-Op EFs | 17.4 | - | 18.2 | 17.7 | - | 17 | 19.2 | 14.7 | 16.1 | 16.9 |
| Mean Drop EFs | 7 | - | 9 | 3.5 | - | 7 | 5.5 | 9.4 | 9.7 | 7.3 |
| Return to Baseline (%) | 39 (24.7) | - | 2 (11.1) | 12 (42.9) | - | 4 (14.8) | 11 (35.5) | 2 (22.2) | 2 (9.5) | 6 (25.0) |
| Statistical Comparison |  |  |  |  |  |  |  |  |  |  |
| Nerve-Sparing |  | Unilateral (p-value) |  |  |  |  | Bilateral (p-value) |  |  |  |
| EF Score Change |  |  |  |  |  |  |  |  |  |  |
| Immediate vs Late |  | +0.6 (1.0) |  |  |  |  | +3.8 (0.04)* |  |  |  |
| Early vs Late |  | +1.6 (0.9) |  |  |  |  | +1.8 (0.9) |  |  |  |
| Immediate vs Early |  | +1 (0.9) |  |  |  |  | +1.9 (0.9) |  |  |  |
| Return to Baseline % |  |  |  |  |  |  |  |  |  |  |
| Immediate vs Late |  | +1.6 (0.6) |  |  |  |  | +17.9 (0.05)* |  |  |  |
| Early vs Late |  | +5.3 (0.5) |  |  |  |  | +10.5 (0.05)* |  |  |  |
| Immediate vs Early |  | -3.7 (0.8) |  |  |  |  | +7.4 (0.1) |  |  |  |

N = Numbers, \* = statistically significant at $p < 0.05$
Uni = Unilateral, Bi = Bilateral, EFs = Erectile Function Score

**Figure. 1.**
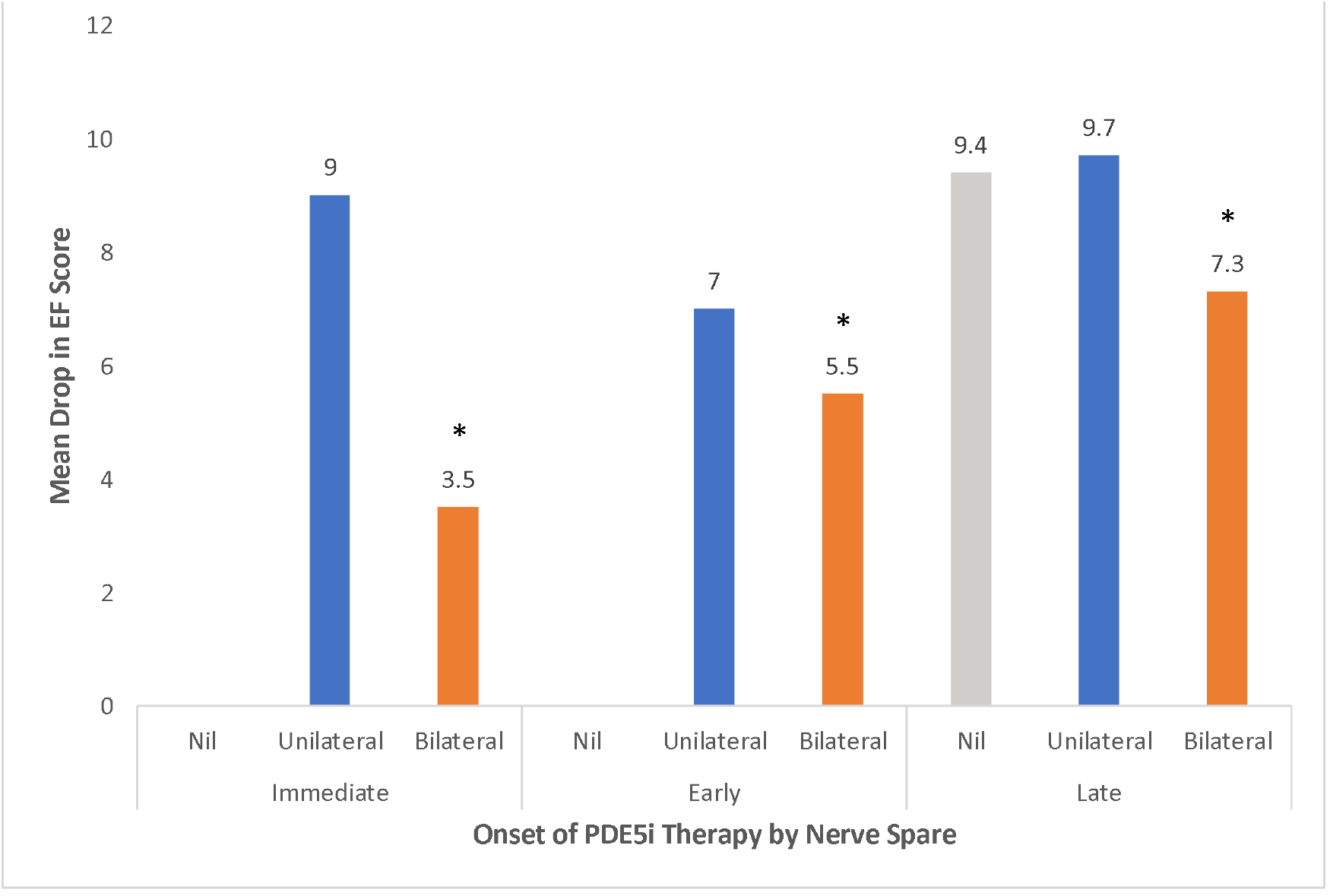
EF Score compared between nerve spare and different PDE5i therapy onset groups. Showing significantly lower drops in EF score with immediate or early therapy in bilateral nerve spare groups. * = statistically significant at p=<0.05

**Figure. 2.**
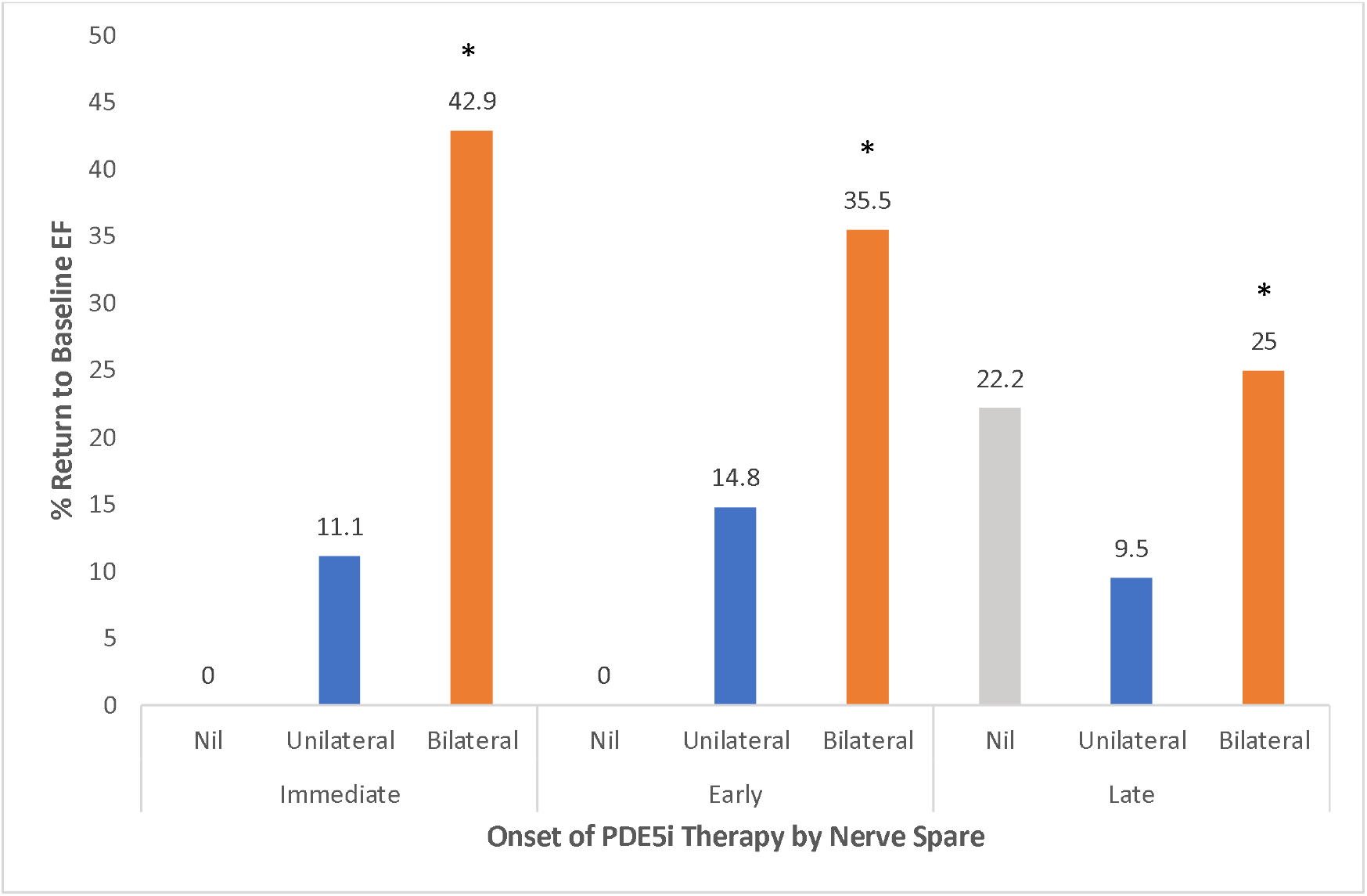
Return to Baseline outcomes compared between nerve spare and different PDE5i therapy onset groups. Showing significantly improved return to baseline EF with immediate or early therapy in bilateral nerve spare groups. * = statistically significant at p=<0.05

### Continence Outcomes

Overall, in our entire cohort, full continence was achieved by 47%, social continence 41%, with 12% remaining incontinent, at a follow-up of 6-8 weeks. Between therapy groups, rates of full and social continence were 54% and 37% in immediate therapy, 60% and 33% in early therapy and 26% and 54% in late therapy (p<0.05) (Table 3) (Fig 3).

**Figure. 3.**
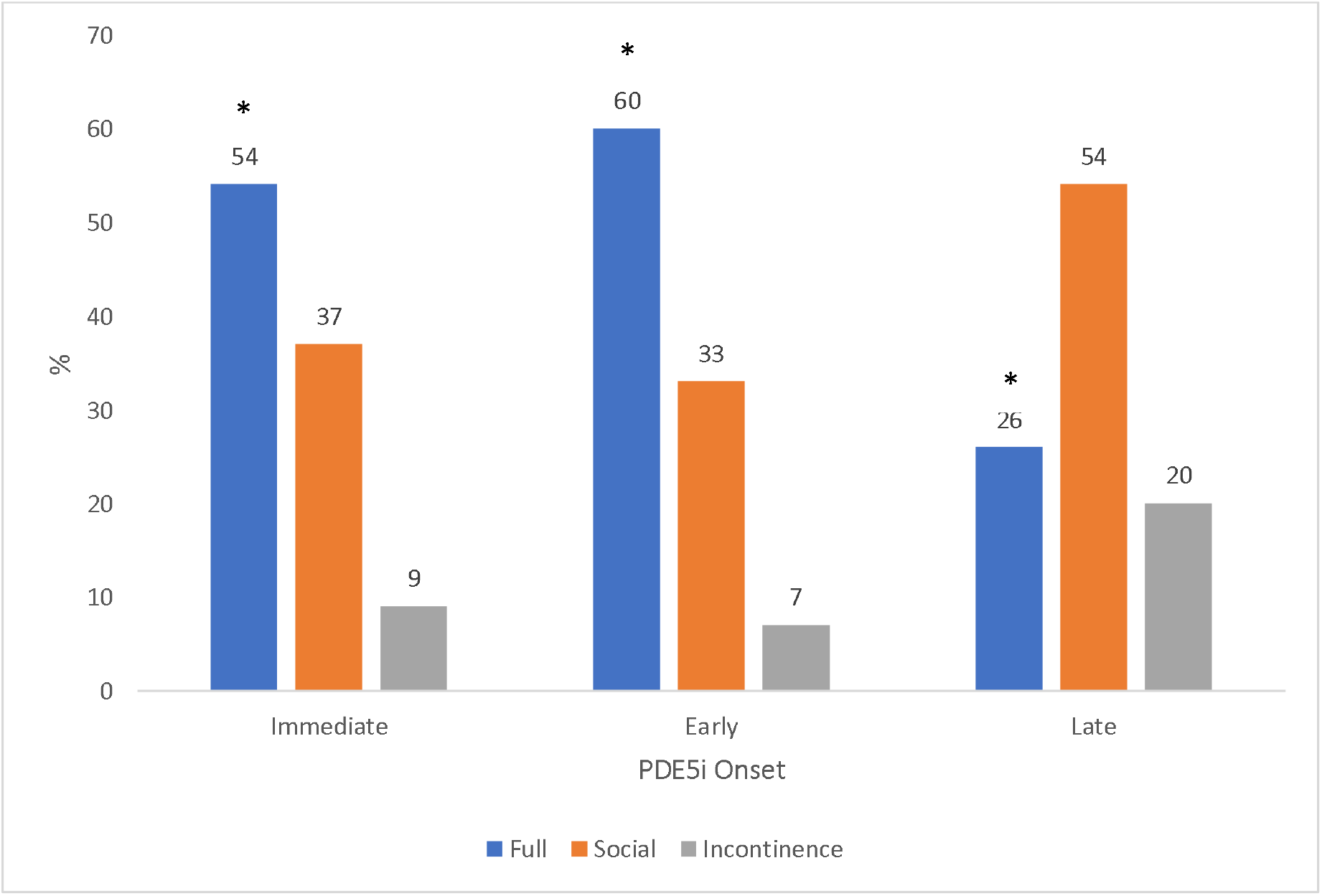
Continence outcomes compared between different PDE5i therapy onset groups. Showing statistically significant improved return to full continence with immediate and early therapy. * = statistically significant at p=<0.05

### Safety Outcomes

PDE5i therapy, in the total cohort, was stopped in 13%, with 7% stopping due to side effects and 6% due to non-compliance. From the entire cohort 2% were readmitted and 5% developed 90-day complications of which one was a haematoma, managed conservatively. There were no statistically significant differences between the three groups for discontinuation, complications or readmission (Table. 3).

## Discussion

Our results suggest that patients who were started earlier on PDE5i therapy after RARP enjoyed better EF outcomes overall. Specifically, the mean drop in EF score showed a trend towards being reduced in the immediate (5.7) and early (6.6) therapy groups compared to late therapy (8.6) group for all nerve sparing groups, though this did not reach statistical significance (p=0.097). Furthermore, return to baseline EF score trended towards improvement in the immediate (36%) and early (39%) groups compared to 26% in late group, though this was not statistically significant (p=0.374).

However, when utilising nerve sparing techniques, immediate therapy showed significant benefit compared to late therapy. These results were more pronounced in bilateral nerve sparing. For bilateral neve sparing, immediate therapy had a higher EF score by 3.8 compared to late therapy (p=0.04). Additionally, return to baseline in immediate therapy was 17.9% higher compared to late therapy (p=0.05) and early therapy was 10.5% higher than late therapy (p=0.05). Immediate and early PDE5i therapy in unilateral nerve sparing also showed greater EF outcomes compared to late therapy, however the differences were not statistically significant.

Continence outcomes were also better in immediate or early therapy compared to late therapy for all patients. Full continence and social continence were achieved by 54% and 37% in the immediate group, 60% and 33% in the early group and only 26% and 54% in the late group (p=0.001). Additionally, there were no differences in compliance, complications or readmissions between the three therapy groups.

A novel finding from our data showed 11% (n=18) of our cohort recovered EF above baseline. This is possibly due to untreated pre-existing erectile dysfunction. In this cohort median pre-operative EF was 11 with a median increase of 4.

Our results show that the level of preservation of the neurovascular bundle is important in erectile function recovery post RARP, as previously described (7, 33). For example, Greco *et al* (34) showed 69% return to sexual intercourse with bilateral nerve sparing compared to 43% in unilateral. Additionally, Sridhar *et al* (21) found that for patients with non-nerve sparing surgery, post-operative treatment with PDE5i made no significant difference to EF.

PDE5i therapy is a well-established component of penile rehabilitation after RARP. Montorsi *et al* (25) showed significantly greater EF scores using on-demand Sildenafil compared to placebo. Pace *et al* (26) also showed improved potency rates using Sildenafil (87%) compared to placebo (56%). Generally, starting PDE5i therapy earlier post-operatively showed better EF score results (35). However, our study adds further weight to the previous limited evidence that earlier post-operative PDE5i therapy improves EF. We showed a mean drop in EF and return to baseline was 5.7 and 36% in the immediate group, 6.6 and 39% in the early group and 8.6 and 26% in the late group. Comparison of different PDE5i has shown 71% of patients preferred Tadalafil and 29% preferred Sildenafil due to side-effect and erection profile (36).

Studies have shown that PDE5i therapy after RARP may also improve urinary continence outcomes (19, 37, 38). Kaiho *et al* (24) showed that immediate PDE5i therapy may temporarily worsen immediate incontinence compared to late therapy. However, our data showed an improvement in early continence rates with immediate PDE5i therapy compared to late therapy. Furthermore, considering anecdotal and pharmacological concerns regarding increased bleeding, our study did not show any differences in adverse effects when initiating PDE5i therapy immediately after RARP (28).

Numerous studies have documented the poor quality of life associated with ED, with most suffering from moderate to severe psychological effects. Common concerns reported included partnership issues, guilt, anger and self-deprecation (39, 40).

Rates of ED after RARP have been reported to be as high as 68% and non-surgical interventions such as focal therapy and radiotherapy report lower rates of ED (7, 41-43). Therefore, immediate PDE5i therapy after RARP should be considered in order to achieve the best possible EF outcome for patients and improve overall satisfaction after RARP.

Despite multiple studies proving the benefit of PDE5i therapy after RARP, there is no clear consensus or pathway implemented for the management of ED after RARP. In general, PDE5i therapy is started four to six weeks post-operatively however our results show that immediate or early PDE5i therapy can improve EF outcomes with no adverse effects. We recommend the use of immediate PDE5i therapy for patients undergoing nerve sparing RARP to improve EF outcomes, continence outcomes and patient satisfaction.

Limitations of this study include single-surgeon data with an associated surgical learning curve confounder which may mask smaller effects in the results. The study also lacks randomisation and multiple follow-up intervals to assess longer term trends.

Further research with randomisation of patients in multiple centres with longer follow-up would improve the quality of the results. Pre-operative PDE5i therapy may also show a benefit to EF outcomes. Additionally, a cost-benefit analysis of extended PDE5i use with respect to quality of life should be undertaken.

## Conclusion

In conclusion, immediate or early post-operative PDE5i therapy after RARP trends towards better early EF outcomes compared to late therapy. This effect is significantly more pronounced in patients undergoing bilateral nerve sparing. There may also be a benefit in EF for patients with unilateral nerve sparing, however the effects are less pronounced. Immediate therapy can also improve early continence outcomes and does not show an increase in adverse outcomes. Preservation of sexual function and regaining continence are important factors for patients undergoing RARP and therefore immediate therapy should be considered especially in bilateral nerve sparing cases.

## Data Availability

Anonymised raw, tabulated data is available

## Acknowledgments

Urology Department Addenbrooke’s Hospital, Cambridge University Hospitals

## References

1. UK CR. Prostate Cancer Incidence Statistics 2019 [Available from: https://www.cancerresearchuk.org/health-professional/cancer-statistics/statistics-by-cancer-type/prostate-cancer/incidence#heading-Zero.

2. Audit NPC. NPCA Annual Report 2019. 2020.

3. Orvieto MA, Patel VR. Evolution of robot-assisted radical prostatectomy. Scand J Surg. 2009;98(2):76–88.

4. Abboudi H, Khan MS, Guru KA, Froghi S, de Win G, Van Poppel H, et al. Learning curves for urological procedures: a systematic review. BJU Int. 2014;114(4):617–29.

5. Krambeck AE, DiMarco DS, Rangel LJ, Bergstralh EJ, Myers RP, Blute ML, et al. Radical prostatectomy for prostatic adenocarcinoma: a matched comparison of open retropubic and robot-assisted techniques. BJU Int. 2009;103(4):448–53.

6. Trinh QD, Sammon J, Sun M, Ravi P, Ghani KR, Bianchi M, et al. Perioperative Outcomes of Robot-Assisted Radical Prostatectomy Compared With Open Radical Prostatectomy: Results From the Nationwide Inpatient Sample. European Urology. 2012;61(4):679–85.

7. Ficarra V, Novara G, Ahlering TE, Costello A, Eastham JA, Graefen M, et al. Systematic Review and Meta-analysis of Studies Reporting Potency Rates After Robot-assisted Radical Prostatectomy. European Urology. 2012;62(3):418–30.

8. Tewari A, Srivasatava A, Menon M. A prospective comparison of radical retropubic and robot-assisted prostatectomy: experience in one institution. BJU Int. 2003;92(3):205–10.

9. Kim SC, Song C, Kim W, Kang T, Park J, Jeong IG, et al. Factors Determining Functional Outcomes After Radical Prostatectomy: Robot-Assisted Versus Retropubic. European Urology. 2011;60(3):413–9.

10. De Groote R, Nathan A, De Bleser E, Pavan N, Sridhar A, Kelly J, et al. Techniques and Outcomes of Salvage Robot-Assisted Radical Prostatectomy (sRARP). Eur Urol. 2020.

11. Huri HZ, Mat Sanusi ND, Razack AH, Mark R. Association of psychological factors, patients’ knowledge, and management among patients with erectile dysfunction. Patient Prefer Adherence. 2016;10:807–23.

12. Latini DM, Penson DF, Colwell HH, Lubeck DP, Mehta SS, Henning JM, et al. Psychological impact of erectile dysfunction: validation of a new health related quality of life measure for patients with erectile dysfunction. J Urol. 2002;168(5):2086–91.

13. Montorsi F, Wilson TG, Rosen RC, Ahlering TE, Artibani W, Carroll PR, et al. Best Practices in Robot-assisted Radical Prostatectomy: Recommendations of the Pasadena Consensus Panel. European Urology. 2012;62(3):368–81.

14. Corbin JD. Mechanisms of action of PDE5 inhibition in erectile dysfunction. Int J Impot Res. 2004;16(1):S4–S7.

15. John Hubert WP. Robotic Urology. 3 ed: Springer International Publishing; 2018.

16. Briganti A, Gallina A, Suardi N, Capitanio U, Tutolo M, Bianchi M, et al. Predicting erectile function recovery after bilateral nerve sparing radical prostatectomy: a proposal of a novel preoperative risk stratification. J Sex Med. 2010;7(7):2521–31.

17. Nelson CJ, Scardino PT, Eastham JA, Mulhall JP. Back to baseline: erectile function recovery after radical prostatectomy from the patients’ perspective. J Sex Med. 2013;10(6):1636–43.

18. Mulhall JP, Burnett AL, Wang R, McVary KT, Moul JW, Bowden CH, et al. A Phase 3, Placebo Controlled Study of the Safety and Efficacy of Avanafil for the Treatment of Erectile Dysfunction After Nerve Sparing Radical Prostatectomy. Journal of Urology. 2013;189(6):2229–36.

19. Patel HR, Ilo D, Shah N, Cuzin B, Chadwick D, Andrianne R, et al. Effects of tadalafil treatment after bilateral nerve-sparing radical prostatectomy: quality of life, psychosocial outcomes, and treatment satisfaction results from a randomized, placebo-controlled phase IV study. BMC Urol. 2015;15:31.

20. Brock G, Montorsi F, Costa P, Shah N, Martinez-Jabaloyas JM, Hammerer P, et al. Effect of Tadalafil Once Daily on Penile Length Loss and Morning Erections in Patients After Bilateral Nerve-sparing Radical Prostatectomy: Results From a Randomized Controlled Trial. Urology. 2015;85(5):1090–6.

21. Sridhar AN, Cathcart PJ, Yap T, Hines J, Nathan S, Briggs TP, et al. Recovery of Baseline Erectile Function in Men Following Radical Prostatectomy for High-Risk Prostate Cancer: A Prospective Analysis Using Validated Measures. J Sex Med. 2016;13(3):435–43.

22. Teloken P, Mesquita G, Montorsi F, Mulhall J. Post-radical prostatectomy pharmacological penile rehabilitation: practice patterns among the international society for sexual medicine practitioners. J Sex Med. 2009;6(7):2032–8.

23. Berkels R, Klotz T, Sticht G, Englemann U, Klaus W. Modulation of human platelet aggregation by the phosphodiesterase type 5 inhibitor sildenafil. J Cardiovasc Pharmacol. 2001;37(4):413–21.

24. Kaiho Y, Yamashita S, Ito A, Kawasaki Y, Izumi H, Kawamorita N, et al. Phosphodiesterase type 5 inhibitor administered immediately after radical prostatectomy temporarily increases the need for incontinence pads, but improves final continence status. Investig Clin Urol. 2016;57(5):357–63.

25. Montorsi F, Brock G, Lee J, Shapiro J, Van Poppel H, Graefen M, et al. Effect of nightly versus on-demand vardenafil on recovery of erectile function in men following bilateral nerve-sparing radical prostatectomy. Eur Urol. 2008;54(4):924–31.

26. Pace G, Del Rosso A, Vicentini C. Penile rehabilitation therapy following radical prostatectomy. Disabil Rehabil. 2010;32(14):1204–8.

27. Jo JK, Jeong SJ, Oh JJ, Lee SW, Lee S, Hong SK, et al. Effect of Starting Penile Rehabilitation with Sildenafil Immediately after Robot-Assisted Laparoscopic Radical Prostatectomy on Erectile Function Recovery: A Prospective Randomized Trial. J Urol. 2018;199(6):1600–6.

28. Schwartz BG, Kloner RA. Drug interactions with phosphodiesterase-5 inhibitors used for the treatment of erectile dysfunction or pulmonary hypertension. Circulation. 2010;122(1):88–95.

29. Wei JT, Dunn RL, Litwin MS, Sandler HM, Sanda MG. Development and validation of the expanded prostate cancer index composite (EPIC) for comprehensive assessment of health-related quality of life in men with prostate cancer. Urology. 2000;56(6):899–905.

30. Tewari AK, Srivastava A, Huang MW, Robinson BD, Shevchuk MM, Durand M, et al. Anatomical grades of nerve sparing: a risk-stratified approach to neural-hammock sparing during robot-assisted radical prostatectomy (RARP). BJU Int. 2011;108(6B):984–92.

31. Sridhar AN, Abozaid M, Rajan P, Sooriakumaran P, Shaw G, Nathan S, et al. Surgical Techniques to Optimize Early Urinary Continence Recovery Post Robot Assisted Radical Prostatectomy for Prostate Cancer. Curr Urol Rep. 2017;18(9):71.

32. Nathan A, Mazzon G, Pavan N, De Groote R, Sridhar A, Nathan S. Management of intractable bladder neck strictures following radical prostatectomy using the Memokath(®)045 stent. J Robot Surg. 2019.

33. Ficarra V, Borghesi M, Suardi N, De Naeyer G, Novara G, Schatteman P, et al. Long-term evaluation of survival, continence and potency (SCP) outcomes after robot-assisted radical prostatectomy (RARP). BJU Int. 2013;112(3):338–45.

34. Greco F, Hoda MR, Wagner S, Reichelt O, Inferrera A, Magno C, et al. Bilateral vs unilateral laparoscopic intrafascial nerve-sparing radical prostatectomy: evaluation of surgical and functional outcomes in 457 patients. BJU Int. 2011;108(4):583–7.

35. Padma-Nathan H, McCullough AR, Levine LA, Lipshultz LI, Siegel R, Montorsi F, et al. Randomized, double-blind, placebo-controlled study of postoperative nightly sildenafil citrate for the prevention of erectile dysfunction after bilateral nerve-sparing radical prostatectomy. Int J Impot Res. 2008;20(5):479–86.

36. Eardley I, Montorsi F, Jackson G, Mirone V, Chan MLS, Loughney K, et al. Factors associated with preference for sildenafil citrate and tadalafil for treating erectile dysfunction in men naive to phosphodiesterase 5 inhibitor therapy: post hoc analysis of data from a multicentre, randomized, open-label, crossover study. BJU Int. 2007;100(1):122–9.

37. Hyndman ME, Bivalacqua TJ, Feng ZY, Mettee LZ, Su LM, Trock BJ, et al. Nightly sildenafil use after radical prostatectomy has adverse effects on urinary convalescence: Results from a randomized trial of nightly vs on-demand dosing regimens. CUAJ-Can Urol Assoc J. 2015;9(11-12):414–9.

38. Gacci M, Ierardi A, Delle Rose A, Tazzioli S, Scapaticci E, Filippi S, et al. Vardenafil can Improve Continence Recovery after Bilateral Nerve Sparing Prostatectomy: Results of a Randomized, Double Blind, Placebo-Controlled Pilot Study. J Sex Med. 2010;7(1):234–43.

39. Korfage IJ, Pluijm S, Roobol M, Dohle GR, Schroder FH, Essink-Bot ML. Erectile Dysfunction and Mental Health in a General Population of Older Men. J Sex Med. 2009;6(2):505–12.

40. Tomlinson JM, Wright D. Impact of erectile dysfunction and its subsequent treatment with sildenafil: qualitative study. BMJ-British Medical Journal. 2004;328(7447):1037–9.

41. Bahn DK, Silverman P, Lee F, Badalament R, Bahn ED, Rewcastle JC. Focal prostate cryoablation: Initial results show cancer control and potency preservation. J Endourol. 2006;20(9):688–92.

42. Ellis DS, Manny TB, Rewcastle JC. Focal cryosurgery followed by penile rehabilitation as primary treatment for localized prostate cancer: Initial results. Urology. 2007;70(6A):9–15.

43. Cozzarini C, Rancati T, Palorini F, Avuzzi B, Garibaldi E, Balestrini D, et al. Patient-reported urinary incontinence after radiotherapy for prostate cancer: Quantifying the dose-effect. Radiother Oncol. 2017;125(1):101–6.

